# Integration of Group A Streptococcus Rapid Tests with the Open Fluidic CandyCollect Device

**DOI:** 10.1101/2024.09.16.24312510

**Authors:** J. Carlos Sanchez, Ingrid H. Robertson, Victoria A. M. Shinkawa, Xiaojing Su, Wan-chen Tu, Timothy R. Robinson, Megan M. Chang, Andrea Blom, Elena Alfaro, Daniel B. Hatchett, Ayokunle O. Olanrewaju, Ellen R. Wald, Gregory P. DeMuri, Erwin Berthier, Sanitta Thongpang, Ashleigh B. Theberge

## Abstract

The CandyCollect device is a lollipop-inspired open fluidic oral sampling device designed to provide a comfortable user sampling experience. We demonstrate that the CandyCollect device can be coupled with a rapid antigen detection test (RADT) kit designed for Group A Streptococcus (GAS). Through *in vitro* experiments with pooled saliva spiked with *Streptococcus pyogenes* we tested various reagents and elution volumes to optimize the RADT readout from CandyCollect device samples. The resulting optimized protocol uses the kit-provided reagents and lateral flow assay (LFA) while replacing the kit’s pharyngeal swab with the CandyCollect device, reducing the elution solution volume, and substituting the tube used for elution to accommodate the CandyCollect device. Positive test results were detected by eye with bacterial concentrations as low as the manufacturer’s “minimal detection limit” - 1.5×10^5^ CFU/mL. LFA strips were also scanned and quantified with image analysis software to determine the signal-to-baseline ratio (SBR) and categorize positive test results without human bias. We tested our optimized protocol for integrating CandyCollect and RADT using CandyCollect clinical samples from pediatric patients (n=6) who were previously diagnosed with GAS pharyngitis via pharyngeal swabs tested with RADT as part of their clinical care. The LFA results of these CandyCollect devices and interspersed negative controls were determined by independent observers, with positive results obtained in four of the six participants on at least one LFA replicate. Taken together, our results show that CandyCollect devices from children with GAS pharyngitis can be tested using LFA rapid tests.

**Table of Contents/Abstract Figure:** 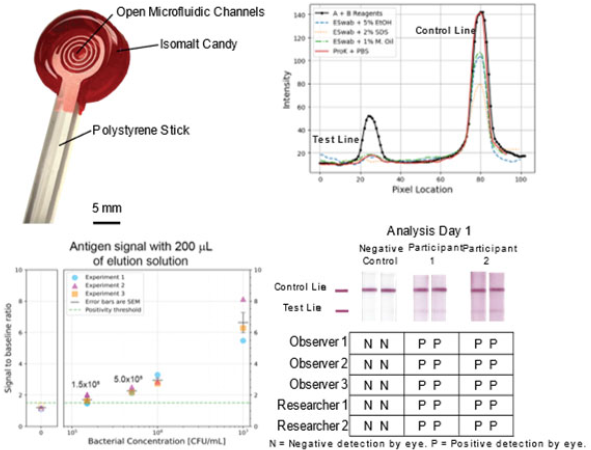

## Introduction

*Streptococcus pyogenes* is responsible for over 600 million cases of bacterial pharyngitis globally each year.^1^ Demographically, this affects 15% of school-aged children and 4-10% of adults; in developing nations, the burden is much greater with rates 5-10 times higher.^1^ Streptococcal pharyngitis most commonly presents as a fever and sore throat an inflamed pharynx and tonsils (often with patchy white exudates). Most symptoms of strep throat resolve after 3 to 4 days, however treatment in the form of antibiotics is recommended to shorten the course of the infection, lessen spread, and prevent more serious outcomes.^1^ Untreated incidents of GAS infections may trigger several disorders including acute rheumatic fever, acute poststreptococcal glomerulonephritis, and pediatric autoimmune neuropsychiatric disorders associated with streptococcal infections. Acute rheumatic fever can lead to long term heart damage known as rheumatic heart disease in children 5-14, resulting in 282,000 new cases each year.^1^

GAS pharyngitis is typically diagnosed by rapid antigen detection test (RADT) which requires a posterior pharyngeal swab sample and provides a positive or negative result usually within 10 minutes. If the RADT is negative, a throat culture is commonly performed requiring laboratory analysis, taking additional time.^2^ Wide-spread usage of RADT has been a valuable tool in detecting strep throat, and has simultaneously reduced the overprescription of antibiotics.^3^ Most importantly, RADTs are inexpensive, offer reasonable sensitivity and specificity, and have short turnaround times.^4,5^ Molecular testing, such as the cobas® liat system (polymerase chain reaction, PCR), is also available requiring a posterior pharyngeal swab and instrumentation.^6^ PCR technology is valuable due to its increased sensitivity at detecting GAS.^7^ Pharyngeal swabs are uncomfortable, sometimes acting as a deterrent for children and adults but it is important to confidently diagnose bacterial pharyngitis not only for the long-term health of the patient but also to avoid over-prescription of antibiotics.^1^

Saliva sampling holds significant promise; saliva is an accessible diagnostic biospecimen for point-of-care devices as it is easy to collect, handle, and test.^8,9^ Here, we present an alternative sample collection device called CandyCollect developed in our lab, ^10,11,12^ and we show for the first time that the CandyCollect device can be used in conjunction with an ‘on the market’ GAS RADT. Inspired by a lollipop, the CandyCollect device is a polystyrene sampling tool that has open fluidic channels on one side and a strawberry-flavored isomalt candy coating on the back and sides. While the user enjoys the candy, pathogens in saliva are collected in the open fluidic channels.^10,11,12^ There are multiple sampling devices currently available that resemble a lollipop but lack some of the built-in features that make the CandyCollect device unique. For instance, Self-LolliSponge™, V-check COVID-19, and Whistling COVID-19 are saliva collection devices that allow the user to self-collect saliva with a minimally invasive protocol, but while these devices resemble a lollipop form they do not contain candy and do not look like a lollipop.^13,14^

Our prior research determined that the open fluidic channel in the CandyCollect device prevents the tongue from removing bacteria, effectively collecting and accumulating the sample for future analysis.^10^ We have previously demonstrated the functionality of the CandyCollect device with an at-home human subjects study that focused on the collection of commensal bacteria for the detection of *Staphylococcus aureus* and *Streptococcus mutans* using qPCR.^11^ We found that the CandyCollect device can successfully capture commensal bacteria in an at-home setting, is stable through standard shipping without refrigeration or other cooling mechanisms, and bacteria can be eluted and quantified using qPCR.^11^ The study also asked users to compare the CandyCollect device to two other commercial methods of collection—a spit tube and ESwab™; users ranked the CandyCollect device as their preferred method of oral sampling. Lastly, it was determined that the CandyCollect device is functional after storage for up to one year.^11^ Subsequently, we tested the CandyCollect device in a clinical setting, enrolling 30 pediatric patients, aged 5-14 years who had positive results from pharyngeal swabs processed with RADT as part of their clinical care.^12^ Results from the CandyCollect device (qPCR analysis) had 100% concordance with the positive results from their clinical care. Further, most children preferred the CandyCollect device over pharyngeal swabs and mouth swabs.^12^

Our prior work used qPCR to detect bacteria (*S. pyogenes, S. mutans, S. aureus*) collected on the CandyCollect devices, which is more sensitive than RADTs but requires specific equipment and is not accessible to a home setting or some clinics. Here, we demonstrate that the CandyCollect device can be integrated with a commercially available RADT as an alternative sample collection tool to the posterior pharyngeal swab commonly packaged with RADT kits. To the best of our knowledge, our work presents the first example of integrating a saliva collection method (the CandyCollect device) with a GAS RADT. In addition, we demonstrate that our integrated protocol can achieve a positive signal on previously frozen clinically sampled CandyCollect devices collected from pediatric patients.^12^

## Material & Methods

### Fabrication of CandyCollect devices

Device fabrication was described in our prior work.^10,11,12^ See SI for full description.

### Capture, elution, and detection of *S. Pyogenes*

#### Capture of S. pyogenes on the CandyCollect device

CandyCollect devices were incubated with 50 μL of *S. pyogenes* suspended in saliva for 10 min with the following concentrations: 1.0×10^9^, 1.0×10^7^, 1.0×10^6^, 5.0×10^5^, 1.5×10^5^ CFU/mL.^1^ Additionally, we incorporated, *S. pyogenes* at 1×10^9^ CFU/mL suspended in THY liquid media as a positive control and filtered pooled saliva as a negative control.

#### Elution of S. pyogenes from the CandyCollect device

Upon completion of the 10 min incubation time, the CandyCollect devices were placed in 14 mL round-bottom polypropylene test tubes (Falcon CAT# 352059) that contained elution reagents, shaken for 15 s, and eluted for another 45 s.

For the majority of experiments, the primary elution reagents used were 2.0 M Sodium Nitrite (Reagent A) and 0.4 M Acetic Acid (Reagent B), obtained from the Areta Strep A Swab Test Kit™ (Easy@Home, CAT# ARST-100S). Elution solutions of 100 µL of Reagents A and B were prepared for each rapid test, resulting in 200 µL total volume. This volume was increased to 200 µL of each reagent for a total of 400 µL of elution solution for our experiment that directly compared our integrated workflow with the kit’s established workflow (per manufacturer’s instructions). The solution was then aliquoted into 14 mL tubes for the CandyCollect device or the kit-provided tubes for the swabs.

Other reagents tested as elution solutions included (1) ESwab™ buffer (Becton, Dickinson and Company, Cat # R723482) with 1% Mineral Oil (Thermo Scientific™, CAT#AC415080010), (2) ESwab™ buffer with 5% ethanol (3) ESwab™ buffer with 2% SDS, and (4) phosphate-buffered saline (PBS) (Fisher Scientific, Cat# BP2944100) with 1% Proteinase K (Thermo Scientific™, Cat# EO0491).^10,11^

#### Clinical sample integration

One of the CandyCollect devices from each replicate set was eluted and analyzed via qPCR following the protocol established in our prior work.^11,12^ The second device was used to test the integrated CandyCollect-Areta™ protocol. Six CandyCollect devices were chosen to test with a requirement that both the replicate CandyCollect and the ESwab™ have low Ct values scores (a CandyCollect was not chosen if the replicate had a low Ct but the ESwab™ did not). It should be noted that we tested the replicate CandyCollect device, not the device that was eluted for qPCR. To prevent inconsistencies with the test strips, all test strips used during a given experiment came from the same box and lot number. For quality assurance, the box was validated per Areta™ new package protocol using the provided negative and positive controls before any tests were started. They were also secondarily checked by the researchers using pooled saliva as a negative control before testing the clinical samples. Clinical sample CandyCollect devices were removed from a -80 °C freezer and thawed on ice for 10 min before testing.

Independent observers, away from the testing area, were given verbal instructions to not discuss results and were given an insert from the Areta™ box that instructs what is considered a negative or positive test result (Figure S1). Negative control devices were inoculated using pooled saliva and were included arbitrarily among the clinical devices. Two strips were tested for each device. At the 18 min mark, the strips were placed inside a sterile OmniTray™ (ThermoScientific™ Nunc™, CAT# 140156) with a clear lid. With the strips inside the tray, independent observers were asked to record whether they thought the test to be negative or positive.

### Analysis of lateral flow immunoassay strips

#### Interpretation of lateral flow immunoassay strip readout by eye

Once the CandyCollect devices were eluted for 1 min, a lateral flow immunoassay strip was immersed into the eluted solution. The immunoassay strip was held in the solution until the solution wicked up the testing area on the immunoassay strip. Upon wicking, a timer was set for 18 min at which point the immunoassay strip was viewed by two researchers and scanned (Hewlett-Packard, HP OfficeJet Pro6978, SN# TH0AK4N0YT). All readouts conducted in the lab were determined by eye from each of the two researchers after 18 min per the manufacturer’s instructions which indicate that results are valid from 10 to 20 min after the test strip is removed from the elution solution. As noted in the Results section, for clinical samples, test strips were viewed by two researchers and three independent observers who are not authors of this publication.

#### Quantification of signal on Areta lateral flow immunoassay strip via optical analysis

The scanned LFA strips were analyzed using a custom image analysis Python script, drawing on algorithms similar to previously published work.^15^ Images of each strip were digitally cropped from an original full-color image obtained by the HP scanner, yielding multiple 25 × 100 pixel images. These cropped images were then converted to monochrome and subsequently inverted. Profiles of digital numbers against pixel location were obtained by averaging the rows for each strip. The script pinpoints the test line peak signal values and locations from the profiles. For each profile, two regions of interest (ROIs) spanning between 5 to 10 pixels were manually selected, approximately 30 pixels from both sides of the peak. These two ROIs, selected from the non-reactive part of the membrane, establish the baseline. The baseline is computed as the average digital number from these two ROIs. The signal-to-baseline ratio (SBR) is calculated as the ratio of the peak signal of the test line from the profile divided by the baseline.

The positivity threshold is a predetermined value of the SBR above which the test results are deemed positive. It is derived from image analysis of the three negative controls collected for each experiment and is calculated from the following equation:

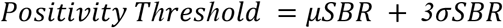

where *μSBR* and *σSBR* are the average signal-to-baseline and the standard deviation signal-to-baseline of the three negative controls, respectively. The python script determines the negative signal by analyzing a segment of the profile located at a specific distance from the negative control line peak, a distance that is informed by the test line peak locations in positive test strips. This approach ensures that the segment location accurately reflects the expected position of a negative control test line.

### Results and Discussion

#### Integration of the CandyCollect device with a commercially available lateral flow immunoassay for GAS detection

The CandyCollect device (Figure 1a.) is a lollipop-inspired oral sampling device that uses plasma-treated open fluidic channels to capture pathogens in saliva as the user consumes the strawberry-flavored isomalt candy.^10,11,12^ We integrated the CandyCollect device into the Areta Strep A Swab Test™ protocol by substituting the kit-provided swab with our novel oral sampling device in order to provide a more comfortable testing experience. There are several commercially available LFA for GAS detection. We explored three commercially available tests, Areta Strep A Swab Test ™, Abbot BinaxNow Strep A Card ™, and Quidel QuickVue Dipstick Strep A Test ™, and compared them based on limit of detection and overall user workflow. The Areta Strep A Swab Test™ kit was selected as the rapid strep throat test to integrate with the CandyCollect device as it had the lowest limit of detection (1.5 × 10^5^ CFU/mL *S. pyogenes*) and the workflow of the test allowed for the most direct substitution of the throat swab sampling method.

**Figure 1.**
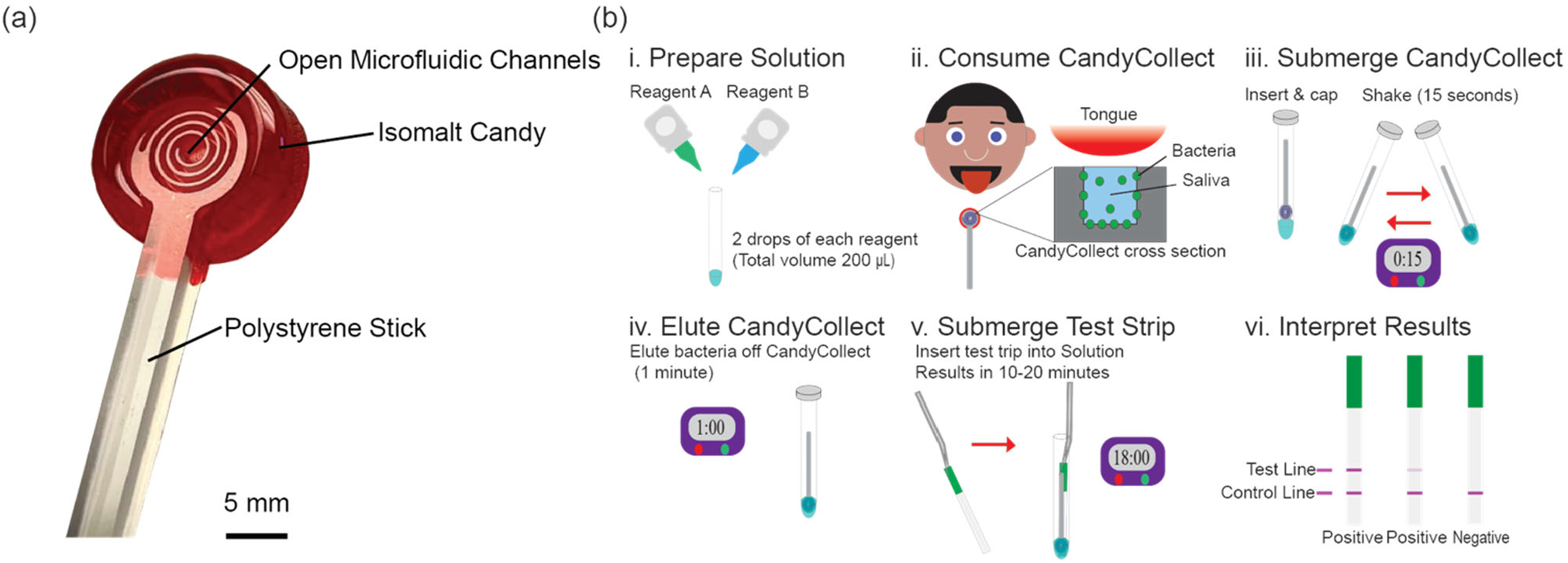
(**a**) Schematic of the CandyCollect Device. The open fluidic channels milled into the head of the plasma-treated polystyrene stick allow pathogens to adhere over sampling time as the user consumes the isomalt candy. (**b**) Workflow to incorporate sampling with the CandyCollect device with the Areta Strep A Swab Test™ protocol. (i) Elution solution is prepared before sampling in the kit-provided tube by combining 100 µL (2 drops) of Reagent A (2.0 M sodium nitrite) and 100 µL (2 drops) of Reagent B (0.4 M acetic acid) for a total elution solution volume of 200 µL. (ii) To sample, the participant places the CandyCollect device in their mouth and consumes the isomalt candy on the CandyCollect device until the candy is completely dissolved. *S. pyogenes* collects on the plasma-treated open-fluidic channels within the CandyCollect device over time. (iii) After sampling, the CandyCollect device is added to the tube of elution solution, and the tube is capped and shaken for 15 s to ensure that the elution solution reaches the channels. (iv) The tube is then set aside for 1 minute. (v) The tube is then uncapped, and the kit-provided test strip is dipped into the elution solution for 10 s or until the fluid visibly starts to wick on the testing section of the lateral flow strip. Test results are valid for 10 to 20 min after complete wicking. (vi) The presence of a test and control line indicates a positive result; a control line only indicates a negative result.

Our optimized integrated workflow is outlined in Figure 1b. Rather than taking a sample directly from the throat with the kit-provided swab, the user’s saliva sample is gathered using the channels of the CandyCollect device. From there, the device is added to a reduced volume of 200 µL of the kit-provided elution reagents [Reagent A (2.0 M sodium nitrite) and Reagent B (0.4 M acetic acid)]. The CandyCollect device is larger than the kit-provided swab, so our protocol includes a larger round-bottom tube to accommodate the device and ensure effective coverage of the CandyCollect channels when immersed in elution solution. The user directly tests the elution solution by dipping the LFA strip following the manufacturer’s instructions. As is common with LFAs, there is a control line and a test line, the presence of both lines indicates a positive result, while the presence of only the control line indicates a negative result. It is important to note that the sample tested in this workflow is saliva rather than a direct collection of bacteria from the user’s throat, like in a throat swab.

#### Optimization of sample preparation and lateral flow assay workflow

We first evaluated the optimal reagents to elute *S. pyogenes* or *S. pyogenes* antigens from the CandyCollect device. We compared the strength of the test line signal when using the kit-provided reagents [Reagent A (2.0 M sodium nitrite) and Reagent B (0.4 M acetic acid)] to the signal strength when using other methods our lab previously developed for eluting *S. pyogenes* prior to qPCR detection (Figure 2).^10^ The elution methods used in the evaluation were ESwab™ buffer with 5% ethanol, ESwab™ buffer with 2% sodium dodecyl sulfate, ESwab™ buffer with 1% mineral oil and PBS with 1% Proteinase K.^10,11^ Test strips were imaged 18 minutes after elution. Quantification of the test line and control line intensity are shown in Figure 2a. Further, results were determined by eye per the manufacturer’s instructions also at 18 minutes after elution and are reported below the corresponding scanned test strip. The scanned images are for visual aid and may not align with the reported results by eye due to quality of imaging.

**Figure 2.**
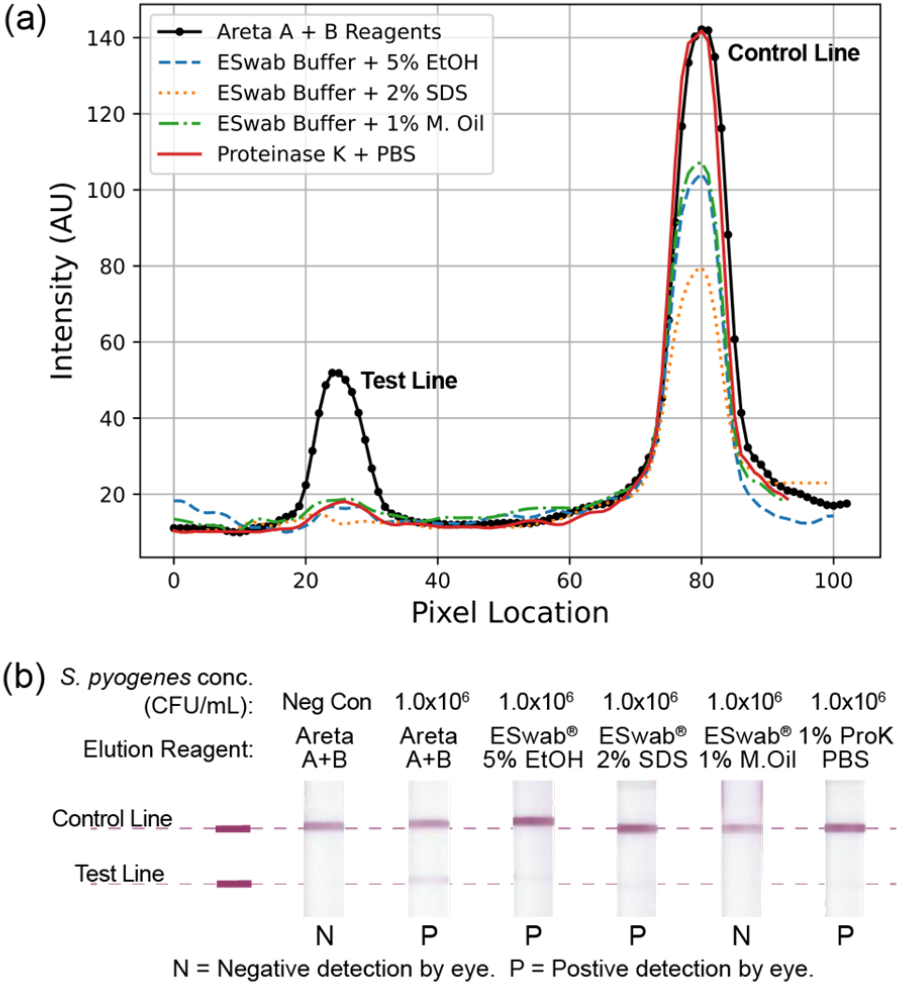
Optimization of elution solution. **(a)** Image analysis of the LFA strips from the elution comparison test shows that the Areta™ kit-provided reagents [Reagent A (2.0 M sodium nitrite) and Reagent B (0.4 M acetic acid)] result in the strongest signal intensity when compared with previous successful elution solutions developed for the CandyCollect device with qPCR analysis.^10,11^ **(b)** Representative images of LFA strips quantified in A (Figure S2 shows images from three independent experiments). *S. pyogenes* at 1.0×10^6^ CFU/mL suspended in filtered pooled saliva was used for all samples. Reagents tested from left to right are: the kit-provided Reagent A (2.0 M sodium nitrite) and Reagent B (0.4 M acetic acid), ESwab™ buffer with 5% ethanol (EtOH), ESwab™ buffer with 2% sodium dodecyl sulfate (SDS), ESwab™ buffer with 1% mineral oil (M. Oil), and phosphate buffered saline (PBS) with 1% Proteinase K (ProK).

The kit-provided reagents resulted in the strongest test line signal across all three experiments (Figure 2 and S2) and were selected as the optimal elution solution for the integrated workflow. ESwab™ buffer with 5% ethanol and PBS with 1% Proteinase K resulted in negative or markedly fainter positive results than the kit-provided reagents. ESwab™ buffer with 2% sodium dodecyl sulfate and ESwab™ buffer with 1% mineral oil consistently resulted in a negative test line, making them unfit as an elution solution for the rapid test kit.

Although the manufacturer’s instructions specify that the strength of the test line signal is not indicative of the concentration of *S. pyogenes*, to aid the user’s ability to read test results easily and accurately, we investigated possible alterations to the rapid test workflow to increase test line signal strength. To increase the concentration of *S. pyogenes* antigens in solution for testing, we reduced the elution solution volume from 400 µL to 200 µL. We investigated the effects of the alteration by directly comparing the rapid test results across a range of *S. pyogenes* concentration samples 1.5×10^5^ to 1.0×10^7^ CFU/mL, with 1.5×10^5^ CFU/mL advertised as the kit’s limit of detection (Figure 3). In order to achieve a non-biased readout, an algorithm was written in Python to evaluate the intensities of the lines, relative to the positivity threshold (Figure S3, S4, S5, & S6). Our intention was to eliminate user discrepancies when interpreting the strips results by eye. The quantified results from three independent experiments are plotted in Figure 3a. Results were also determined by eye per the manufacturer’s instructions 18 minutes after testing the elution solution and are reported below the corresponding scanned test strip (Figure 3b). The scanned images are for visual aid and may not align with the reported results due to quality of imaging.

**Figure 3.**
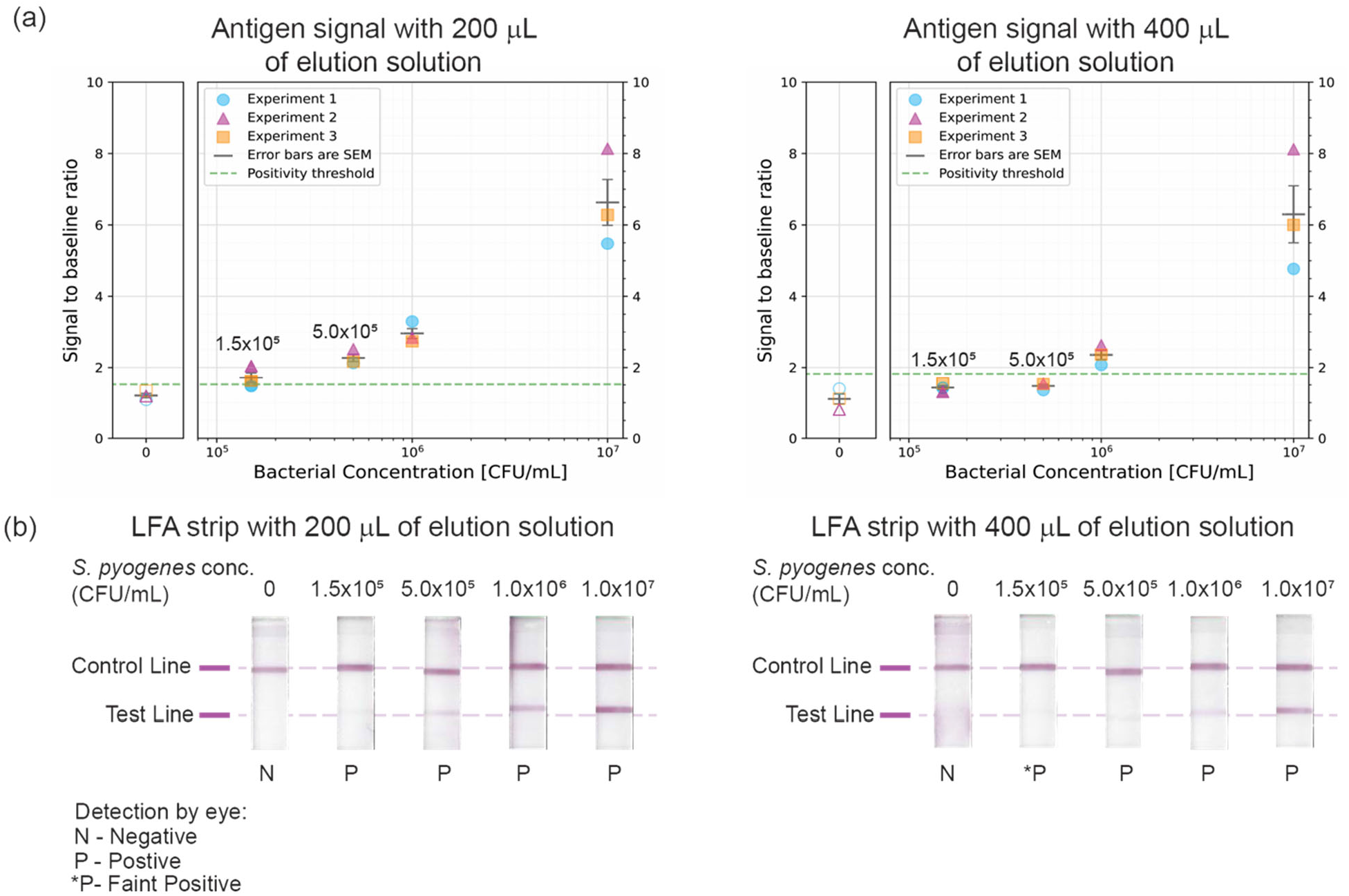
Reduction of elution volume results in a stronger test line signal. Areta Strep A Swab Test™ results using different elution reagent volumes, 200 µL (our optimized reduced volume) and the 400 µL (the volume specified in the Areta test kit instructions). **(a)** Quantification of three independent experiments with the mean and standard error of the mean (SEM). The positivity threshold (green dashed line) is calculated as the mean + three standard deviations of the signal-to-baseline ratio (SBR) taken from the negative controls (Figure S5 & S6). **(b)** Representative images of LFA strips quantified in A (Figure S3 shows images from three independent experiments). Elution reagents used were Reagent A (2.0 M sodium nitrite) and Reagent B (0.4 M acetic acid) provided in the rapid test kit. The different elution volumes were tested using samples across a range of concentrations from 1.5×10^5^ to 1.0 × 10^7^ S. pyogenes CFU/mL.

When testing samples at 1.5 × 10^5^ CFU/mL, the 400 µL elution solution volume resulted in very faintly positive to negative results by eye, however, the reduced 200 µL elution solution volume resulted in positive results across three independent experiments (Figure 3b). Image quantification revealed that for 1.5×10^5^ CFU/mL, the 200 µL elution solution volume resulted in a signal that was above or at the positivity threshold, whereas using the 400 µL elution solution volume resulted in a signal that was below the positivity threshold (Figure 3a). This trend was further confirmed with the 5.0×10^5^ CFU/mL samples, as the strength of the test line signal by eye from the samples using the 200 µL elution solution volume was consistently greater than the test line signal of the samples using the 400 µL elution solution volume; quantification of the test line signal intensity of the 5.0×10^5^ CFU/mL samples showed that using the 200 µL elution solution volume resulted in a signal that was above the positivity threshold whereas using the 400 µL elution solution volume resulted in a signal that was below the positivity threshold (Figure 3a). For 1.0×10^6^ and 1.0×10^7^ CFU/mL samples, both conditions resulted in strong positive results and the difference in strength of signal was less prominent based on image quantification and by eye (Figure 3a and b). Taken together, the results showed the benefit of reducing the elution solution volume to 200 µL, and this change was implemented into our integrated workflow (Figure 1b).

#### CandyCollect sampling method gives comparable results to the kit-provided swab based on in vitro experiments in spiked saliva

To compare the efficacy of the CandyCollect device as an alternative sampling method to the kit-provided swab, we evaluated the strength of the test lines across three different sampling methods and the two different elution solution volumes (Figure 4). For our experiments, we used a 50 µL sample of *S. pyogenes* in saliva to inoculate the channels of the CandyCollect device for 10 minutes as an *in vitro* simulation of bacterial collection from saliva. To compare to the CandyCollect device, we deposited samples onto the kit-provided swab in two ways: (i) 50 µL of the sample directly pipetted onto the swab (to directly compare the same sample volume that was pipetted on the CandyCollect device) and (ii) dipping the swab directly into the saliva sample. An important limitation of the comparisons between swabs and the CandyCollect device is that the swab data from the manufacturer is completed at the back of the throat whereas in this work we used bacteria suspended saliva; however, we decided to compare the sampling methods this way due to practicality of the experimental setup.

**Figure 4.**
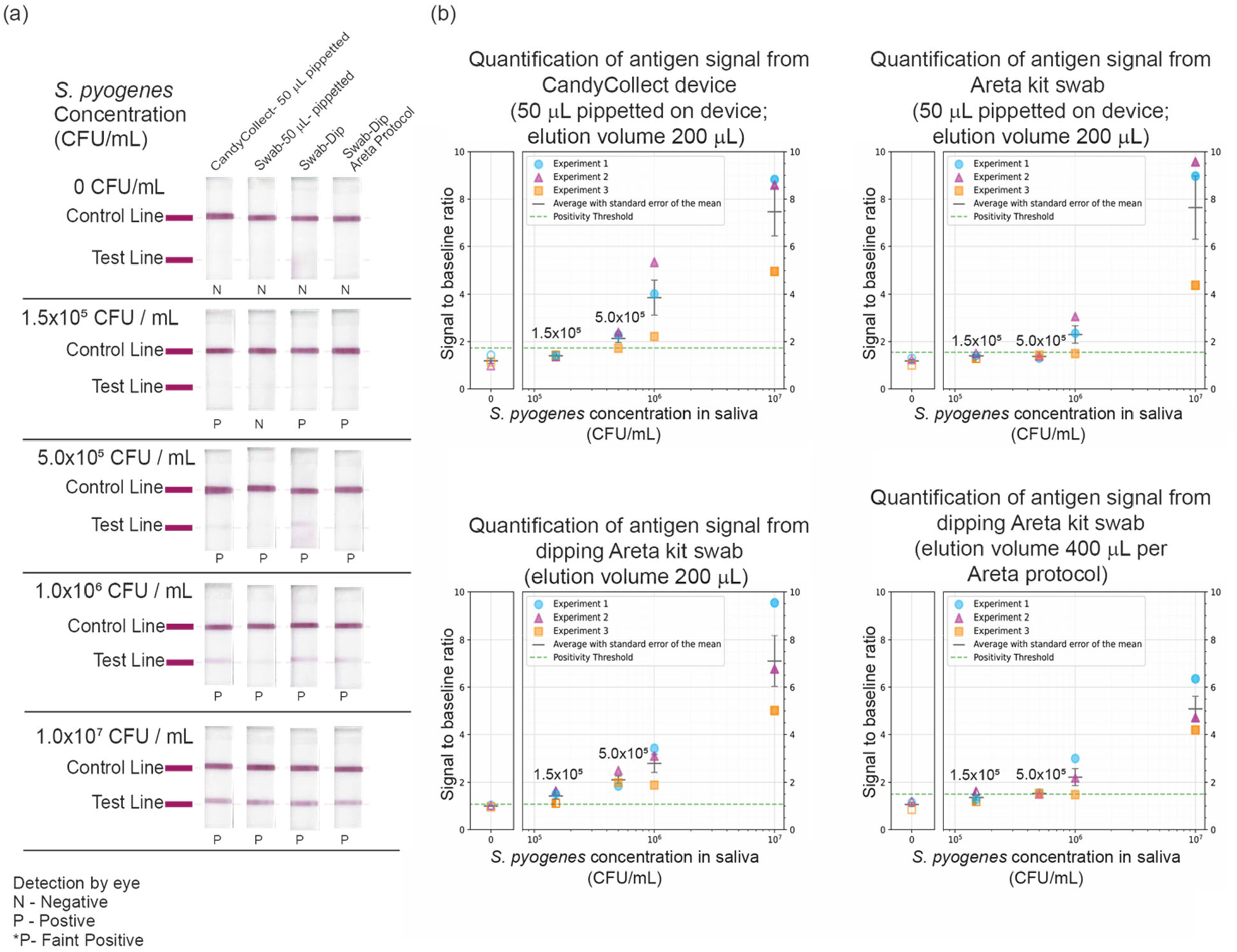
The CandyCollect device gives comparable results to kit-provided swabs. **(a)** Areta Strep A Swab Test™ results using three different sampling methods with the optimized (reduced elution solution volume) workflow (CandyCollect – 50 µL pipetted; kit-provided swab – 50 µL pipetted; and kit-provided swab – dipped into sample all eluted in 200 µL) as well as the original Areta workflow (kit-provided swab – dipped into sample eluted in 400 µL). All four methods were tested across a range of concentrations from 1.5×10^5^ to 1.0×10^7^ CFU/mL *S. pyogenes* in pooled human saliva. The images were taken at 18-20 min, and the result indicated (positive or negative) was determined by eye according to the test instructions at 18 min. Images shown are representative of three independent experiments. **(b)** Quantification of three independent experiments with the mean and standard error of the mean (SEM). The positivity threshold (green dashed line) is calculated as the mean + three standard deviations of the signal to baseline ratio (SBR) taken from the negative controls (see SI for further details).

All four methods were tested across a range of concentrations from 1.5×10^5^ to 1.0×10^7^ CFU/mL *S. pyogenes* suspended in filtered pooled human saliva, as well as a negative control of only saliva. For this experiment, we followed our optimized integrated workflow with a 200 µL elution volume for the three different methods. We also tested a condition using the kit directed workflow with a 400 µL elution volume with the swab dipped into the saliva sample. Both the 50 µL 1.5×10^5^ CFU/mL sample on the CandyCollect device and the dipped samples on the swab gave positive results, however, the 50 µL sample pipetted on the swab produced a negative result by eye (Figure 4a). Image quantification revealed that pipetting 50 µL of 1.5×10^5^ CFU/ml S. *pyogenes* and following up with 200 µL of elution solution onto the CandyCollect device, resulted in a signal below that positivity threshold. Image quantification for the Areta kit swab, using the same conditions, demonstrated a similar result, being at or just below the positivity threshold (Figure 4b). At 5×10^5^ CFU/mL *S. pyogenes* all four methods resulted in positive tests that were visible by eye (Figure 4a). As the image quantification shows, two out of the three methods in this experiment, which used a 200 µL elution volume, resulted in a signal above the positivity threshold line. In contrast, the Areta kit swab, following Areta’s recommended elution volume (400 µL), resulted in a signal at the positivity threshold line (Figure 4b). At 1.0×10^6^ and 1.0×10^7^ CFU/mL *S. pyogenes* all four conditions resulted in positive tests. The results were consistent across all three experiments, specifically that the CandyCollect device sample at 1.5×10^5^ CFU/mL *S. pyogenes* was read as positive by eye, further supporting it as a possible alternative sampling method (Figure 4a). Lastly, at 1.0×10^6^ and 1.0×10^7^ mean values for all conditions were above or at the positivity threshold (Figure 4b).

Of note, there were minor differences between the results in Figures 3a (left plot) and 4b (top left plot) using the same experimental conditions (50 μL of *S. pyogenes* in saliva pipetted onto the CandyCollect device and eluted in 200 μL). The difference across these two sets of experiments is likely due to manufacturing inconsistencies across LFA strips and varying amounts of leaching dye that results in a higher background and therefore higher positivity threshold in Figure 4b than in Figure 3a.

#### Clinical sample results

Subsequently, we tested the CandyCollect device in a clinical setting, enrolling 30 pediatric patients, aged 5-14 years who had positive results from pharyngeal swabs processed with RADT as part of their clinical care.^12^ We performed the optimized protocol (shown in Figure 1) on six CandyCollect devices from six participants. We performed the LFA in duplicate for each of the CandyCollect device (the 200 µL elution volume allowed for two LFA strips). We asked three independent observers who are not co-authors on this manuscript to evaluate LFA strips from negative controls and human subject samples shown to them in an arbitrary order. Two researchers, who are co-authors on this manuscript, also viewed the test strips, and all results are reported in Figure 5. The human subject samples were tested over the course of three days, with fresh negative controls interspersed each day. Independent observers were shown the Areta™ protocol and were told that a line, even a faint line, is positive and no line is negative, as instructed by the Areta™ protocol (see Figure S8). We requested that the independent observers not discuss or share their answers with each other.

**Figure 5.**
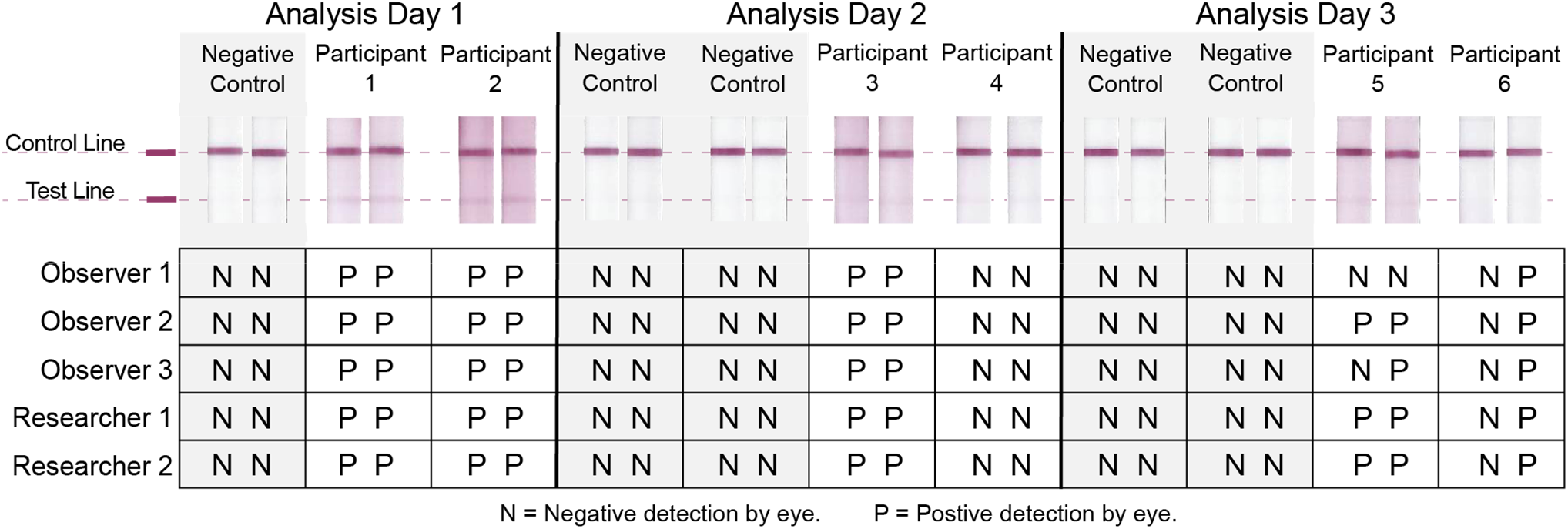
Rapid test results from the CandyCollect device samples obtained from children with GAS pharyngitis. Testing was completed over the course of three days. Participants 1-3 had positive test results on two replicate LFA strips eluted from the sample CandyCollect device. For Participant 6, one LFA strip was positive and the duplicate LFA strip was negative. The LFA strips for participant 5 did not have universal agreement among independent observers and researchers. Independent observers were shown LFA strips in an arbitrary order. The scanned images are for visual aid and may not align with the reported results due to quality of imaging.

Additional red pigment due to leftover candy was common on the devices from the clinical samples, resulting in background pinkish/red color along the length of the test strip, particularly in Participants 1, 2, 3, and 5. In the future we plan to test other candy colors, such as yellow, to avoid this background signal. Further, due to the high background, we did not quantify these images as the human eye is able to discern a positive signal despite the background and the Areta™ protocol instructions call for reading the strips by eye. Twenty out of 22 test strips had unanimous agreement as being either positive or negative; the two that did not have agreement among the independent observers came from the same CandyCollect device (from Participant 5) on analysis day 3; this CandyCollect device, still had candy on the device which caused the testing area to be pinkish/red. The combination of the added pigment and the possibility of the concentration of *S. pyogenes* being near the minimal detection limit for this sample may have led to the mixed observations. Participants 1-3 had positive test results on two replicate LFA strips eluted from the sample CandyCollect device. For Participant 6, one LFA strip was positive and the duplicate LFA strip was negative. Taken together, our results show that the CandyCollect devices from children with GAS pharyngitis can be tested using LFA rapid tests. Further work is needed to reduce the red background (by changing the candy color), to expand the sample size, and to improve detection in samples with less abundant bacteria. We note that the Areta Strep A Swab Test ™ was designed to be integrated with a pharyngeal swab, which will likely have different levels of bacteria compared to a saliva sample; further development of the CandyCollect device surface properties to be specific to capturing GAS may improve the levels of bacteria collected and is a subject of future work.

## Conclusion

In this work, we combined a previously reported novel saliva sampling device with an available GAS RADT. This was achieved by altering the Areta™ kit protocol: (1) replacing the pharyngeal swab with the CandyCollect device, (2) substituting the kit test tube for a larger round-bottom tube (to accommodate the larger CandyCollect device), and (3) adjusting the elution volume. Other elution solutions were considered but results showed that elution reagents provided by the manufacturer performed best. *In vitro* experiments showed we can consistently achieve a positive test strip signal with the “minimal detection limit” indicated by the manufacturer. Furthermore, previously frozen CandyCollect clinical samples from pediatric patients diagnosed as positive for GAS via RADT performed on pharyngeal swabs as part of their clinical care also yielded a positive signal while following our optimized protocol. Future research will be conducted remotely with children and their parents to continue to test usability and practicality of performing our CandyCollect-rapid test protocol in a home setting. Future studies will also be done to examine other bacterial and viral pathogens found in saliva that can be captured by the CandyCollect device and integrated with rapid tests, such as SARS-CoV-2. We will also continue our ongoing collaborative studies in clinical settings to build our sample size with patients and to further integrate their feedback.

## Data Availability

All data produced in the present work are contained in the manuscript

## Author Contributions

J.C.S., I.R., and V.A.M.S., contributed equally to this work. J.C.S., I.R., V.A.M.S., X.S., E.B.., S.T., and A.B.T conceptualized the research integrating the CandyCollect with RADTs. A.B. E.A., E.R.W., G.P.D., E.B., S.T., and A.B.T. conceptualized the pediatric clinical study, and E.R.W. and G.P.D. oversaw study execution. J.C.S. and I.R. milled the CandyCollect devices. J.C.S., I.R., V.A.M.S., and D.B.H fabricated candy devices and prepared devices for the human subjects research, including designing experimental protocols/engineering designs. J.C.S., I.R., and V.A.M.S. X.S., W.-C.T., reviewed the literature that informed the study, designed the biological experiments, conducted biological experiments and data collection. A.B. advised on study design and execution, enrolled and interacted with the participants, collected the samples and user surveys, and accessioned and shipped the samples. E.A. advised on study design and execution, advised on all regulatory aspects, drafted the study protocol and survey instruments, and obtained Institutional Review Board (IRB) approval. J.C.S., T.R.R., M.M.C., A.O.O., E.B., and A.B.T. developed and refined data analysis methods for quantifying RADT results. All authors analyzed and interpreted the data. J.C.S., I.R., V.A.M.S., X.S., T.R.R., M.M.C., S.T., and A.B.T., wrote sections of the manuscript. All authors revised and approved the manuscript.

## Acknowledgement

All phases of this study were supported by National Institutes of Health grants (R21AI166120, R35GM128648 (the latter specifically supported some of the in-lab developments and procedure developments)), the Washington Research Foundation, a STEP grant from UW CoMotion, the Camille and Henry Dreyfus Foundation, and an Alfred P. Sloan Research Fellowship. The content is solely the responsibility of the authors and does not necessarily represent the official views of the National Institutes of Health or other funding sources. We would like to thank the participants in the clinical study and the independent observers.

