## Supplemental Materials for "Integration of Group A Streptococcus Rapid Tests with the Open Fluidic CandyCollect Device"

**Affiliations:**
[1]Department of Chemistry, University of Washington, Seattle, WA, USA
[2]Department of Mechanical Engineering, University of Washington, Seattle WA, USA
[3]Department of Bioengineering, University of Washington, Seattle WA, USA
[4]Department of Pediatrics, University of Wisconsin School of Medicine and Public Health, Madison, WI, USA
[5]Department of Biomedical Engineering, Faculty of Engineering, Mahidol University, Nakorn Pathom, Thailand
[6]Department of Urology, School of Medicine, University of Washington, Seattle, WA, USA

*These authors contributed equally to this work
[§]Co-corresponding authors

**Address correspondence to:** Ashleigh B. Theberge, Department of Chemistry, University of Washington, Box 351700, Seattle, WA 98195-1700, [], 206-685-2330. Sanitta Thongpang, Department of Chemistry, University of Washington, Box 351700, Seattle, WA 98195-1700, [], 206-685-2330.

**Figure S1:** Areta™ Strep A Swab Test instructing how to read LFA results
**Figure S2:** Images of three independent elution solution experiments
**Figure S3:** Images of three independent elution volume experiment
**Figure S4:** Image of test data used for data analysis
**Figure S5:** Data preparation for the image analysis
**Figure S6:** An example of a signal profile for a strip exposed to a high bacteria concentration
**Figure S7:** An example of a signal profile for a negative control
**Figure S8.** Images of the Instruction Manual that is provided to the user
**Figure S9.** Engineering drawing of CandyCollect device used in in-lab experiments (Figures 2-4)
**Figure S10.** Engineering drawing of CandyCollect device used in human subjects (Figure 5)
**Extended Materials and Methods**

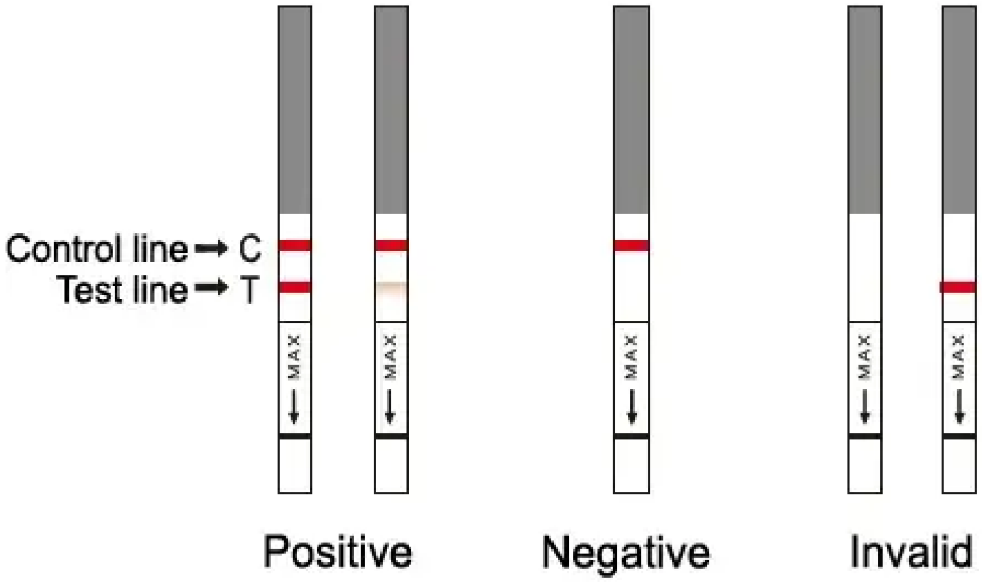


**Figure S1.** Image from insert included in Areta™ Strep A Swab Test instructing how to read LFA results. This was shown to independent observers prior to reviewing test strips for the clinical samples.

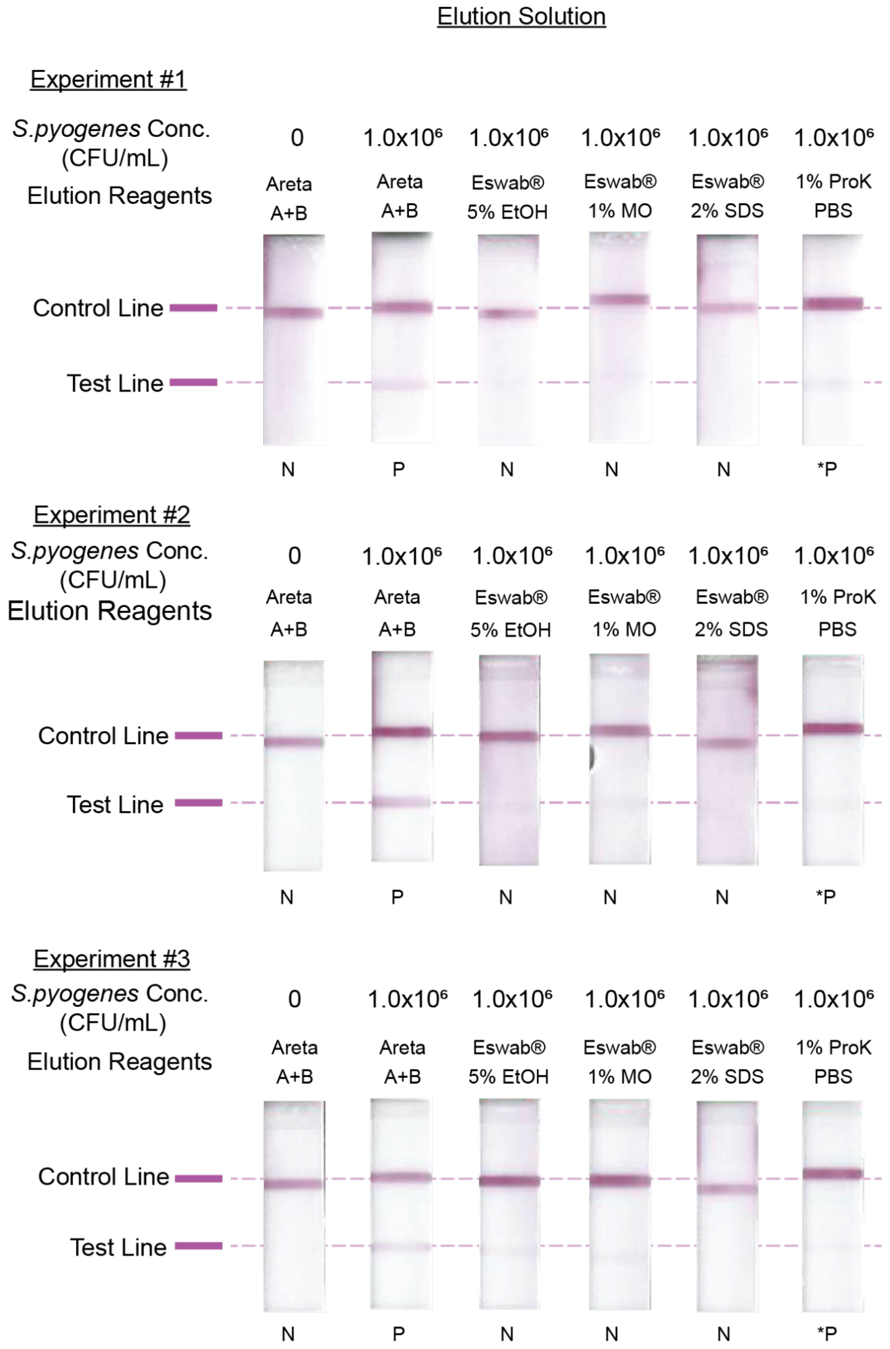


**Figure S2**. Images of the LFA strips from three independent experiments conducted to determine the optimal elution solutions for eluting antigen from the surface of *S. pyogenes* captured on our CandyCollect device. These images support our finding that Areta Reagents A (2.0 M sodium nitrite) and Reagent B (0.4 M acetic acid) are the ideal elution reagents for our experiments. These reagents consistently produced positive results with *S. pyogenes* at a concentration of $1.0 \times 10^6$ CFU/mL in pooled human saliva. Additionally, we tested other potential elution solutions previously used to strip *S. pyogenes* from the surface of the CandyCollect device from our prior work.[1,2] However, the Areta reagents were found to be the most effective in consistently producing positive signals on the LFA strips.

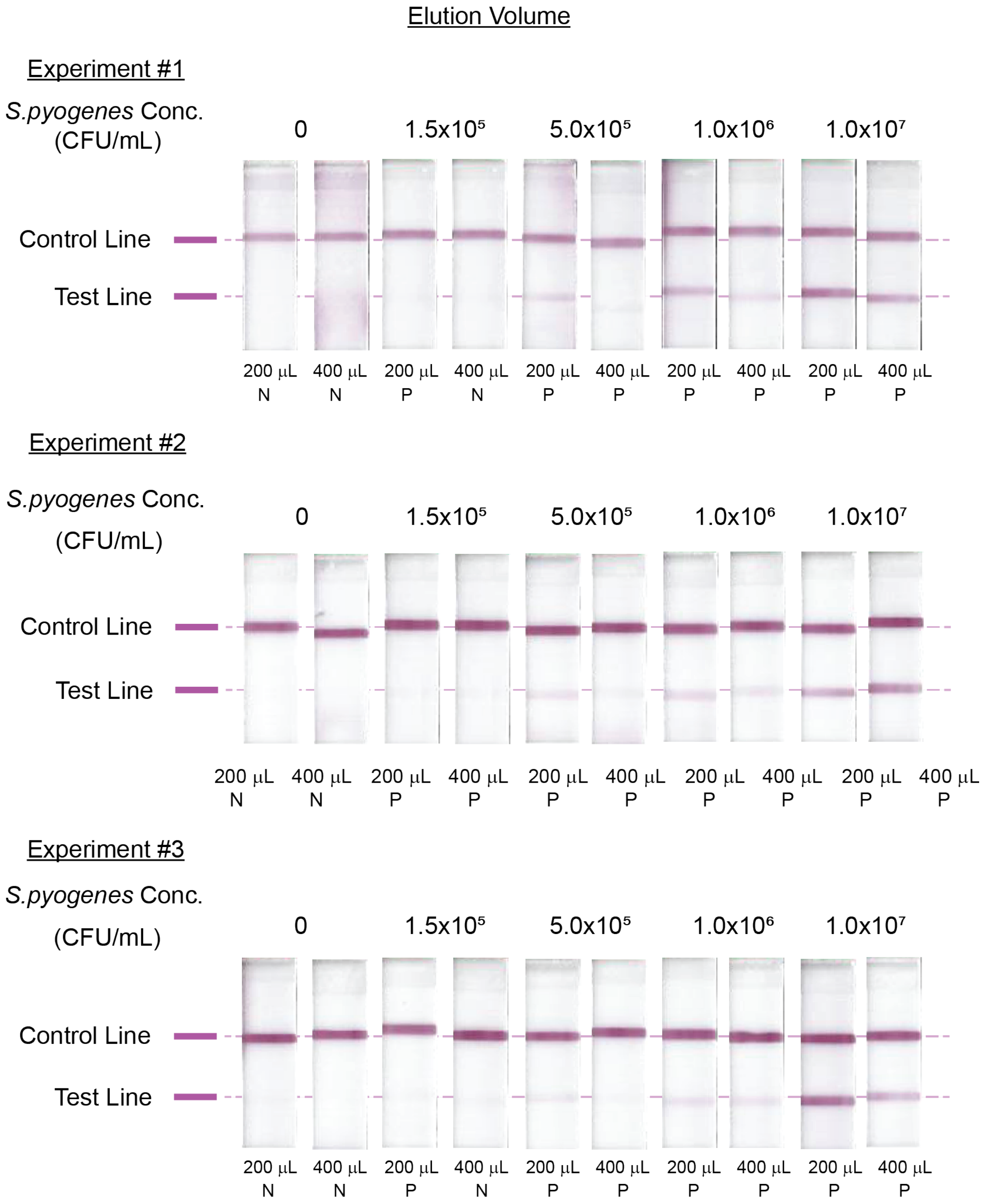


**Figure S3**. Images of the LFA strips of three independent experiments conducted to test our proposed volume alteration. These images support our findings that using 200 µL elution volume provides enhanced signal compared to the prescribed 400 µL from the Areta protocol. In most cases, we did observe stronger signals with 200 µL compared to 400 µL on multiple occasions, and more importantly, at lower *S. pyogenes* concentration.

**Image analysis and quantification**

The signal-to-baseline ratio (SBR) and positivity thresholds, depicted in Figures 2, 3, and 4, were derived using custom image analysis software written in Python version 3.8.16. The data pipeline consists of three subsections: Data Preparation, Determining the Signal-to-Baseline ratios, and Determining Positivity Thresholds.

(1) Data Preparation

Figure S4 shows an example of the original data, which is a full color image (.jpg) of a set of test strips acquired from the flatbed scanner.

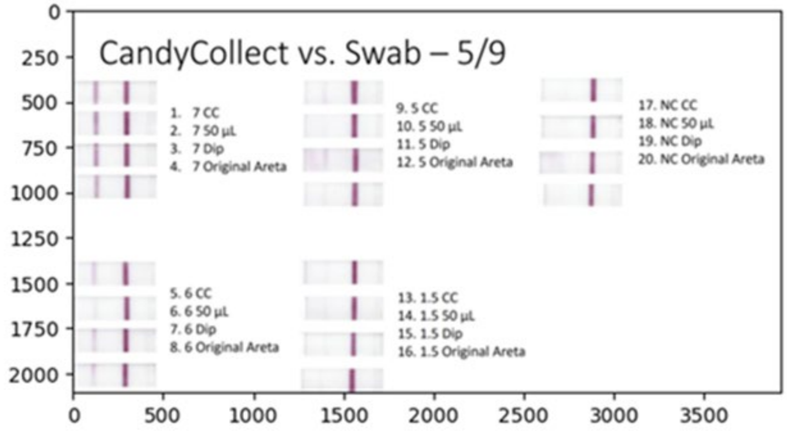


**Figure S4**. Image of test data used for data analysis. The conditions are distinguished as CC (CandyCollect), 50 µL (Pipette onto Swab), Dip (Swab was dipped), and Original Areta. The number to the left of the description indicates the bacteria concentration. The number to the left of the bacteria concentration is an index. As such, there are four different conditions, each tested under 4 different bacteria concentrations. The strips to the far right are the negative controls for each condition.

The software code performs several operations. First, it converts the file to a lossless format (.tiff). All subsequent modifications and analysis are performed on this lossless formatted image file. Next, the code crops each test strip. The full-color (3D array) image is then converted to a monochrome (2D array) image. This monochrome image is then inverted. The inverted image array is normalized to the bit-depth of the scanner imaging system. In our case, the scanner produced a .jpg with an 8-bit depth for each of the color channels. Therefore, the inverted image is normalized by dividing the array by 255.

The analysis revealed that the green channel provided the highest contrast. As such, the monochrome image is derived from the cropped full-color image. Figure S5 shows the results cropping, isolating the green channel isolation, and inverting the image.

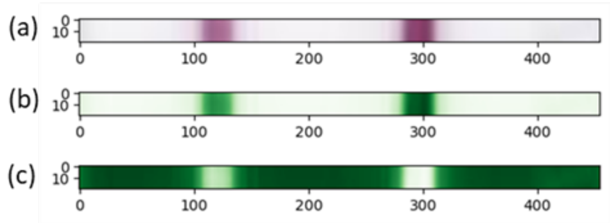


**Figure S5**. Data preparation for the image analysis. (a) A cropped full color image of a test strip. (b) A monochrome image of the test strip (green channel from the full color image). (c) The inversion of the monochrome image.

(2) Determining signal-to-baseline ratios

The profile, representing the digital number signal vs. pixel location, is derived from the cropped and inverted monochrome image. As shown in Figure S4, each cropped image contains 20 rows of data. The profile is generated by averaging these rows. Within this average profile, the code pinpoints the location and value of the test line peak signal.

The baseline is determined by averaging the signals collected from two profile segments. The segments are located approximately ± 80 pixels from the test peak. Occasionally, test strip membranes had a smeared appearance, which may result in profiles with irregular features along the baseline. In this case, the precise segment location is manually selected. Each segment is 30 pixels wide.

The SBR is the ratio of the test line peak signal to the baseline. This procedure is repeated for each strip shown in the original data image.

Figure S6 represents a typical profile from a test strip exposed to a high bacteria load. The red rectangles pinpoint the locations of the two segments used to calculate the baseline. In this example, the test line peak signal intensity is 0.57, and the baseline is 0.05. The SBR is therefore 0.57 divided by 0.05.

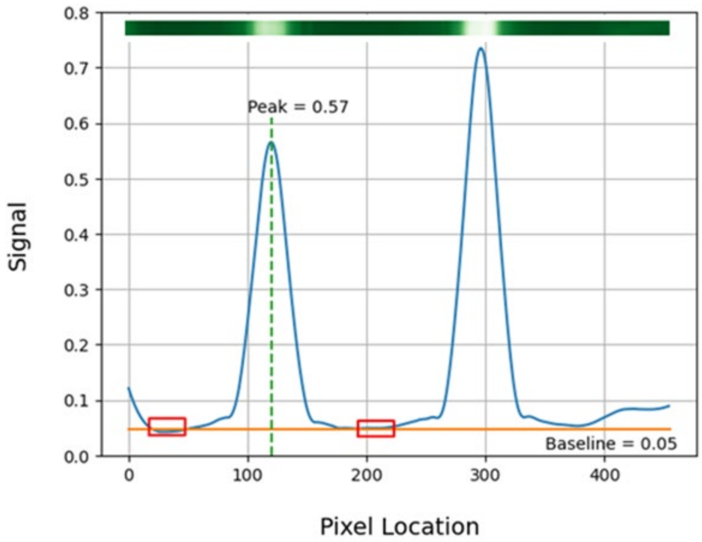


**Figure S6**. An example of a signal profile for a strip exposed to a high bacteria concentration. The left peak identified with the dashed line is from the test line. The peak to the right is from the control line. The red rectangles are auto selected segments used to determine the baseline.

(3) Determining the Positivity Thresholds

Each experimental condition was measured three times, yielding three negative controls. The positivity thresholds for each condition are based on these negative controls.

The first step is to calculate the SBR for the negative controls. However, a problem arises as the negative controls lack a discernible peak in the test line. In contrast, positive test strips consistently show both a test line peak and a control line peak separated by approximately 180 pixels. Consequently, the code identifies a peak signal within a range of 160 to 200 pixels from the control line peak. The baseline is determined as described in the 'Determining signal-to-baseline ratio' section.

Figure S7 represents a typical profile of a negative control. The red rectangles mark the locations of the two segments used to calculate the baseline. In this case, the test line peak signal intensity and baseline are 0.02. The SBR is 0.02 divided by 0.02.

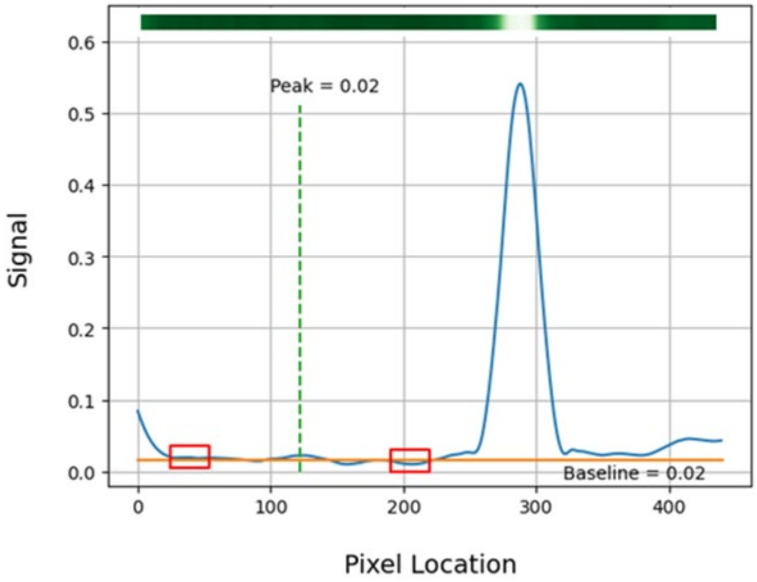


**Figure S7**. An example of a signal profile for a negative control. The signal identified with the dashed line is from the location that approximates the test line. The red rectangles are auto selected segments used to determine the baseline.

The positivity threshold is defined as three-sigma from the average negative control SBRs in accordance to the following equation:

$$\text{Positivity Threshold} = \mu SBR + 3\sigma SBR$$

where $\mu$ symbolizes the mean and $\sigma$ denotes the standard deviation.

### Areta®

#### Strep A Swab Test

Cat. No.: ARST-100S

##### INTENDED USE

Areta Strep A Swab Test is a chromatographic immunoassay for the qualitative detection of Strep A antigen from throat swab specimens of symptomatic patients to aid in the diagnosis of Group A Streptococcal Infection. All negative test results should be confirmed by bacterial culture because negative results do not preclude Group A Strep infection and should not be used as the sole basis for treatment.

For *in vitro* diagnostic use.

##### SUMMARY

Beta-hemolytic group A streptococcus is a common cause of upper respiratory infection in humans, most commonly resulting in pharyngitis. The highest rate of infection is found in children. The infection can lead to serious complications including rheumatic fever and acute glomerulonephritis. Rapid diagnosis and appropriate antibiotic therapy appear to be the best means of preventing these complications. The traditional means of detecting group A streptococcal infection involves 24-48 hour culture of throat swab specimens or other exudates, confirming beta-hemolysis, and showing susceptibility to bacitracin. Accurate rapid diagnosis aids physicians in administering the correct therapy. Areta Strep A Swab Test only needs 12 minutes to get the result after collecting the specimen.

##### PRINCIPLE

Areta Strep A Swab Test utilizes double antibodies sandwich immunoassay for the detection of Group A Streptococcal antigen. The test strip has been pre-coated with rabbit anti- Strep A antibody on the test region and goat anti-rabbit antibody on the control region. When the strip is immersed into the specimen, the specimen is absorbed into the strip by capillary action, mixes with the antibody-dye conjugate, and flows across the pre-coated membrane.

When the group A streptococcal antigen level in the specimens is at or above the target cutoff (the detection limit of the test), the antigen binds to the antibody-dye conjugate and is captured by rabbit anti- Strep A antibody immobilized in the test region of the strip. This produces the colored band and indicates the positive result.

When the group A streptococcal antigen levels are zero or below the target cut off, there is no visible colored band in the test region of the strip. This indicates the negative result.

To serve as a procedure control, a colored line will appear at the control region if the test has been performed properly.

##### PRECAUTIONS

1. This kit is for *in vitro* diagnostic use only. Do not swallow.
2. Do not interchange contents of different lot of kits.
3. Do not interchange caps of reagents.
4. Do not interchange caps of control solution bottles.
5. Do not use the test kit beyond the expiration date.
6. Do not use the test kit if the pouch is punctured or not well sealed.
7. Discard after use. The test kit cannot be used more than once.
8. The extraction tube and swab are for single use. Do not re-use.
9. Reagent A and B are caustic. Avoid contact with eyes, sensitive mucous membranes, cuts, and abrasions etc. If these reagents come in contact with the skin or eyes, flush with a large volume of water.
10. The control solutions contain sodium azide, which, on contact with lead or copper plumbing, may react to form explosive metal azides. Use the large volume of water to flush reagents on disposal.
11. Do not eat, drink or smoke in the area where the specimens and kits are handled.
12. All specimens should be treated as potentially infectious diseases specimens. Protection glove should be worn when handling the specimen. Wash hands thoroughly afterwards.
13. DISPOSAL OF THE DIAGNOSTIC: The used strip, swab and extraction tube have infectious risk. The process of disposing the diagnostic must follow the local infectious disposal law or laboratory rule.

##### MATERIAL

**Material Provided**

1. Each pouch contains one test strip and one desiccant (The desiccant is for storage purpose only, and should not be used in the test procedure.)
2. One extraction tube per test
3. One throat swab per test
4. One extraction reagent A (7mL): 2.0 M sodium nitrite solution (Warning: R25 Toxic if swallowed)
5. One extraction reagent B (7mL): 0.4 M acetic acid solution
6. Two standard controls
   - Positive Control (1mL): Extracted (non-infective) group A streptococcus antigen in phosphate buffer containing 0.1% NaN3. (Warning: R22 Harmful if swallowed)
   - Negative Control (1mL): Phosphate buffer containing 0.1% NaN3. (Warning: R22 Harmful if swallowed)
7. One instruction for use
8. One procedure card

**Material Required But Not Provided**

- Timer

##### STORAGE AND STABILITY

1. Store at 40°F~86°F (4°C ~ 30°C) in the sealed pouch up to the expiration date.
2. Keep away from sunlight, moisture and heat.
3. DO NOT FREEZE.

##### SPECIMEN COLLECTION AND PREPARATION

1. Collect the throat swab specimen with the throat swab that is provided in the kit. Transport swabs containing modified Stuart's or Amies medium can also be used with this product. Swab the posterior pharynx, tonsils and other inflamed areas. Avoid touching the tongue, cheeks and teeth with the swab.
2. Testing should ideally be performed immediately after the specimens have been collected. Swab specimens may be stored at room temperature for up to four hours prior to testing. Note: A second swab may be collected for bacterial culture. Culture should only be conducted by laboratories that are appropriately certified and in accordance with established procedures and practices.

Procedure: If a single swab is collected, culture may be performed first by lightly rolling the swab tip onto a Group A selective (GAS) blood agar plate before using the swab for Areta Strep A Swab Test.

##### TEST PROCEDURE

Allow the test strip and extraction reagents to equilibrate to room temperature 10°C ~30°C(50°F ~86°F) prior to testing.

1. Hold the Reagent A bottle upright and add 4 full drops (approximately 200 μL) to an extraction test tube. Hold the Reagent B bottle upright and add 4 full drops (approximately 200 μL) to the tube. Tap the bottom of the tube gently to mix the liquid.
2. Place the specimen throat swab into the tube. Swirl the swab for 10 times. Leave the swab in the tube for 1 minute. Then remove the swab while squeezing the swab tip against the inside of the extraction tube as remove it to expunge as much liquid as possible from the swab. Discard the swab.
   Mix contents of the tube by gently swirling. **The extraction specimen must be tested immediately.**
3. Remove the test strip from the sealed foil pouch by tearing at the notch. Immerse the strip into the extraction tube with the arrow pointing towards the specimen.
   **IMPORTANT:** Do not allow the specimen level to exceed the MAX Marker Line, otherwise the test will not perform correctly.
   Take the strip out after at least 5 seconds and lay the strip flat on a clean, dry, non-absorbent surface.
4. Wait for 10 minutes and read the results. Do not read results after 20 minutes.

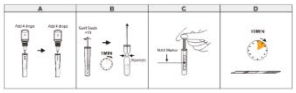


##### INTERPRETATION OF RESULTS

**Positive (+)**
Colored bands are visible in both the control region and the test region. It indicates the positive result for Strep A antigen.

**Negative (-)**
A colored band is visible only in the control region. No color band appears in the test region. It indicates that the concentration of strep A antigen in specimen tested is zero or below the detection limit of the test.

**Invalid**
No visible band at all, or there is a visible band only in the test region but not in the control region. Repeat with a new test strip. If test still fails, please contact the distributor for technical assistance.

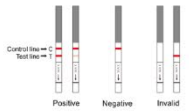


**Note:** There is no meaning attributed to line color, intensity, or width.

##### QUALITY CONTROL

**Internal Procedural Control**

There is an internal procedural control line built in the strip. The appearance of this control band verifies that the test strip is intact and that the sufficient volume of specimen has migrated to the test reaction area. The internal control does not ensure that the strip is working correctly with patient specimen.

**External Positive and Negative Controls**

Good laboratory practice suggests the use of positive and negative controls to ensure that test reagents are working and the test is correctly performed, including the antigen extraction. Areta Strep A Swab Test Kit contains 1 positive control and 1 negative control. Run these controls:
- with each new test kit opened
- with each new operator

The positive control will produce the moderate positive result (two bands- one in the test region and the other in the control region) when the test has been performed correctly and the test strip is functioning properly. The negative control will yield the negative result (one band in control region only) when the test has been performed correctly and the test strip is functioning properly.

**Procedure for External Quality Control Testing**

Allow the strip, extraction reagents and controls to equilibrate to room temperature 50°F~86°F (10°C ~ 30°C) prior to testing.
1. Add 4 drops of extraction reagent A and 4 drops of extraction reagent B respectively into the extraction tube and fully mix.
2. After thoroughly mixing the control, add 3 drops of positive or negative control into this tube. Mix contents by gently swirling.
3. Continue with step 3 to step 5 of **Test Procedure**.

The use of positive and negative controls from other commercial strep A test kits has not been validated with Areta Strep A Swab Test.

##### QUALITY CONTROL

**Internal Procedural Control**

There is an internal procedural control line built in this device. The appearance of this control line verifies that the test device is intact and that a sufficient volume of sample has migrated to the test reaction area. The internal control does not ensure that the device is working correctly with patient samples.

**External Positive and Negative Controls**

Good laboratory practice suggests the use of positive and negative controls to ensure that test reagents are working and the test is correctly performed, including the antigen extraction. Areta Strep A Swab Test kit contains 1 Positive control and 1 Negative control. The Controls will monitor the entire assay. Run these controls:
- with each new test kit opened;
- with each new operator

The Positive control will produce a moderate positive result (two lines—one at the Test region (T) and the other at the Control region (C)) when the test has been performed correctly and the test device is functioning properly. The Negative control will yield a negative result (Control line only) when the test has been performed correctly and the test device is functioning properly.

**Procedure for External Quality Control Testing**

Allow the device, extraction reagents and controls to equilibrate to room temperature 50ºF~ 86ºF (10ºC ~ 30 ºC) prior to testing.
1. Add 4 drops of extraction reagent A and 4 drops of extraction reagent B respectively into the extraction tube and fully mix.
2. After thoroughly mixing the control, add 3 drops of positive or negative control into this tube. Mix contents by gently swirling.
3. Continue with step 3 to step 5 of **Test Procedure.**

The use of positive and negative controls from other commercial kits has not been established with Areta Strep A Swab Test.

##### LIMITATIONS OF PROCEDURE

1. This test has been developed for testing throat swab specimen only. The performance of this test using other specimens has not been substantiated.
2. This test is a qualitative screening assay. It is not designed to determine the quantitative concentration of group A streptococcal antigen.
3. Pharyngitis can be caused by organisms other than group A streptococcus. This test does not provide any further information about pharyngitis other than the possibility of Strep A infection. If clinical signs and symptoms are not consistent with laboratory results, a follow-up throat culture and grouping procedure should be performed.
4. Proper throat swabs must be obtained for good quality tests.
5. A negative result may be obtained due to poor sample collection, or at the onset of the disease due to a low antigen level below the sensitivity limit of the test. If symptoms persist or intensify, retesting with a fresh sample is recommended.
6. A negative result may be obtained due to poor sample collection, or at the onset of the disease due to a low antigen level, below the sensitivity limit of the test. If symptoms persist or intensify, repeat testing with a fresh sample is recommended.
7. As with all diagnostic tests, all results must be interpreted together with other clinical information available to the physician.

##### EXPECTED VALUES

Group A streptococcus infection exhibits a seasonal variation and is most prevalent in the winter and early spring. Approximately 19% of all upper respiratory tract infections are caused by group A streptococcus. The highest incidence of this disease is found in high density populations, such as school aged children and military bases. Males and females are equally affected by the disease .

##### PERFORMANCE CHARACTERISTICS

**Accuracy**

The clinical performance of the Areta Strep A Swab Test was established in a multicenter, prospective Clinical study conducted in 2011～2012 at six geographically diverse physician office, clinics, and emergency departments within the United States. A total of 349 throat swab specimens were evaluated by comparing the Areta Strep A Swab Test to culture method. Of the 349 total specimens, 248 were found to be negative by culture and 101 were found to be positive by culture.

| Results of Areta Strep A Swab Test | Culture Results | | |
|---|---|---|---|
| | Positive | Negative | Total |
| Positive | 96 | 4 | 100 |
| Negative | 5 | 244 | 249 |
| Total | 101 | 248 | 349 |

**Sensitivity**: (96/101) **95.0%** (95% confidence interval: 88.9~97.9%)
**Specificity**: (244/248) **98.4%** (95% confidence interval: 95.9%~99.4%)

**Cross Reactivity**

To confirm the analytical specificity (cross-reactivity) of Areta Strep A Swab Test, organisms likely to be found in the respiratory tract, as listed below, were tested at 1 x $10^8$ organisms per test and were all found to be negative when tested with the Areta Strep A Swab Test.

| | | |
|---|---|---|
| Streptococcus Group B | Candida albicans | Neisseria meningitides |
| Streptococcus Group C | Corynebacterium diphtheriae | Neisseria mucosa |
| Streptococcus Group F | Enterococcus faecalis | Neisseria sicca |
| Streptococcus Group G | Enterococcus faecium | Neisseria subflava |
| Streptococcus salivarius | Escherichia coli | Proteus vulgaris |
| Streptococcus anginosus | Fusobacterium necrophorum | Pseudomonas aeruginosa |
| Streptococcus mitis | Haemophilus parahaemolyticus | Serratia marcescens |
| Streptococcus mutans | Haemophilus parainfluenzae | Staphylococcus marcescens |
| Streptococcus oralis | Haemophilus influenzae | Staphylococcus aureus |
| Streptococcus pneumoniae | Klebsiella pneumoniae | Staphylococcus epidermidis |
| Streptococcus sanguis | Moraxella catarrhalis | Staphylococcus haemolyticus |
| Arcanobacterium haemolyticum | Moraxella lacunata | Yersinia enterocolitica |
| Bordetella pertussis | Neisseria gonorrhoeae | |
| Branhamella catarrhalis | Neisseria lactamica | |

**Analytical Sensitivity**

The minimum detection limit of the test is 1.5 x $10^5$ organisms/mL. This was established by testing inactivated Streptococcus pyogenes with a known number of organisms, ATCC 20159. The organisms were serially diluted and tested by Areta Strep A Swab Test.

**Reproducibility Study**

To investigate the reproducibility of Areta Strep A Swab Test, three lots of tests were utilized in this evaluation. This study was conducted two runs/day on 5 different days at three different sites by testing 4 blind samples. The 4 samples consisted of a true negative samples (diluent only), a moderate positive sample (2.3×$10^6$ organisms /mL), a cut-off sample (1.5×$10^5$ organisms /mL, C95 concentration, approximately positive 95% of the time), and a low negative sample (0.4x$10^5$ organisms /mL) . Six experienced professional operators who didn't know the sample number code participated in the study, for two operators at each testing site. Each operator tested two runs per day at each concentration sample with three lots of Areta Strep A Swab Test. A total of 30 determinations by each operator, at each concentration, were made. There was no significant difference to test results of Areta Strep A Swab Test by different users in different sites on different days.


##### INDEX OF SYMBOLS

| | | | | | |
|---|---|---|---|---|---|
| | See instruction for use | Σ | Tests per kit | | Manufacturing date |
| IVD | For *in vitro* diagnostic use only | | Expiry date | | Do not reuse |
| 30°C / 4°C | Store between 39º~86ºF (4º~30ºC) | LOT | Batch number | REF | Catalog # |
| | Keep away from sunlight | | Keep dry | | |

**Figure S8.** Images of the Instruction Manual that is provided to the user.

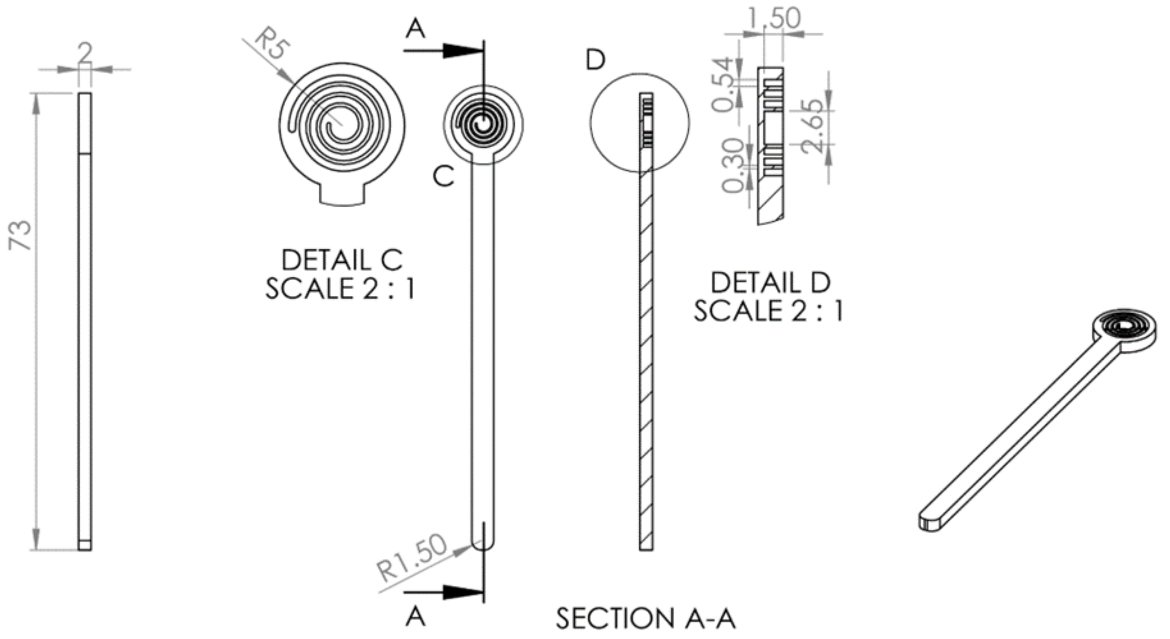


**Figure S9.** Engineering drawing of CandyCollect device used in in-lab experiments (Figures 2-4). Reproduced from Lee et al.[1] All dimensions are in mm.

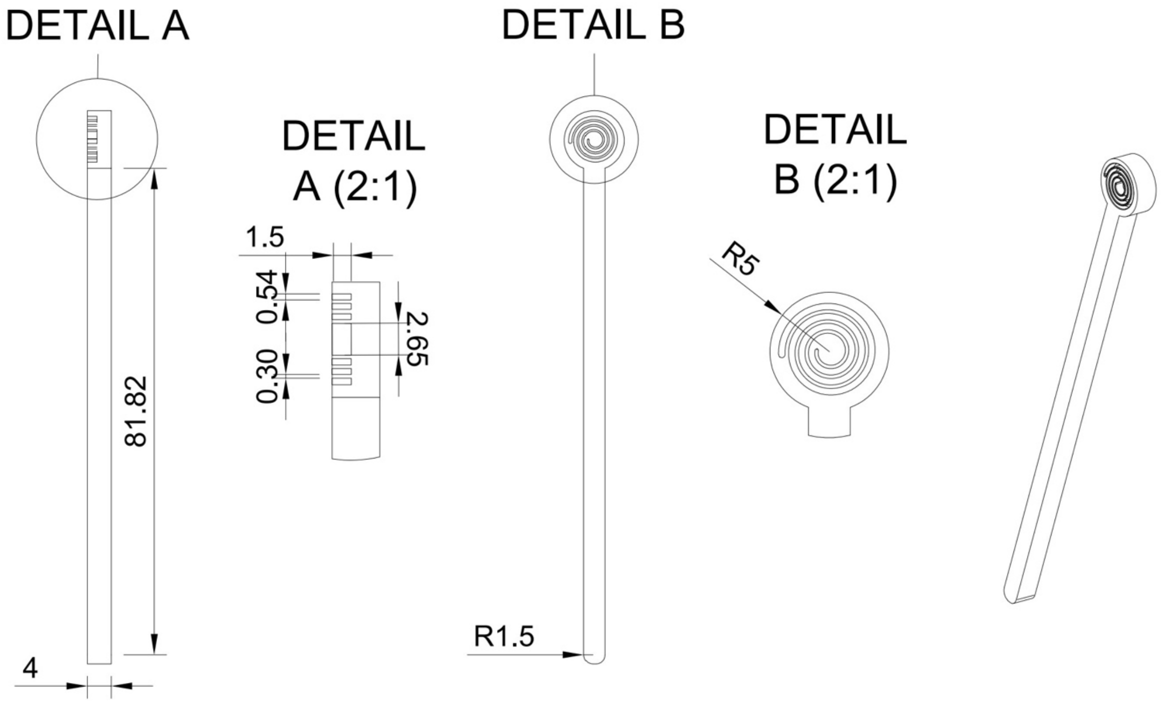


**Figure S10.** Engineering drawing of CandyCollect device used in human subjects (Figure 5). Reproduced from Tu et al.[2] All dimensions are in mm.

#### Extended Materials and Methods

##### Fabrication of CandyCollect devices

*CandyCollect device milling*

Using a computer numerical control (CNC) DATRON neo milling machine, the in-lab experimental CandyCollect devices were milled from 2 mm thick polystyrene sheets (GoodFellow, Cat# 235-756-86) Afterwards, the CandyCollect devices were sonicated with isopropanol (IPA) (FisherScientific, A451-4) and 70% v/v ethanol (FisherScientific, Decon™ Labs, 07-678-004).[1,2]

*CandyCollect device surface activation*

CandyCollect devices were surface activated by plasma treating the devices with oxygen using a Zepto LC PC Plasma Treater (Diener Electronic GmbH Plasma Treater, EbHausen, Germany). The following protocol is consistent with our previous publications.[1,2] In brief, atmospheric gas present in the chamber was removed to achieve a pressure of about 0.20 mbar. Afterwards, the chamber was backfilled with oxygen gas to establish a pressure of 0.25 mbar. Next, the chamber was exposed to 70 W of voltage for 5 min.[1,2]

##### Bacteria culture

*Culture of* S. pyogenes *in Todd-Hewitt yeast (THY) broth*

Experiments were carried out using *Streptococcus pyogenes* strain SF 370 (American Type Culture Collection, ATCC®, Cat#700294). *S. pyogenes* was cultured in Todd-Hewitt Broth (TH Broth) (BD Bacto ™ TH broth, Fisher Scientific, Cat#DF0492-17-6) containing 0.2% yeast extract (United States Biological Corporation, FischerScientific, Cat# NC9796728)(THY) or THY agar plate.[1,2,3,4] *S. pyogenes* was inoculated from a frozen stock, which is stored in a -80 °C freezer, into a 15 mL conical tube (Fisher Scientific, Falcon ™, Cat# 14-959-49B) containing 10 mL of THY liquid media. It was then cultured overnight in a 37 °C incubator supplied with 5% $CO_2$.

*Maintenance of* S. pyogenes *with THY agar*
To maintain the bacteria, *S. pyogenes* were streaked on agar plates with a sterile disposable inoculating loop (Globe Scientific, Fisher Scientific, Cat# 22-170-201). The agar plates were incubated at 37 °C with 5% $CO_2$ overnight, then stored at room temperature for up to 7 days. The day before the experiments, a single colony from an agar plate was used to inoculate 10 mL of THY liquid media in a 15 mL conical tube (Fisher Scientific, Falcon ™, Cat# 14-959-49B) and incubated overnight as above.[1,2,3,4]

*Preparation of* S. pyogenes *in filtered pooled human saliva*
Optical density at 600 nm (OD600) was measured to estimate the concentration of *S. pyogenes* growing in liquid culture media using a Visible 721-Vis Spectrophotometer (Vinmax). Liquid culture was centrifuged to pellet cells for 5 min at 10000 rpm. The *S. pyogenes* pellet was resuspended in filtered pooled human saliva (Innovative Research, Cat# IRHUSL50ML; filtered using a 0.22 µm filter). To achieve desired concentrations of *S. pyogenes*, serial dilutions were performed. See the section below for OD600 to CFU/mL conversion.

*Converting between OD600 absorbance measurements of* S. pyogenes *to CFU/mL*
To generate OD600 to CFU/mL conversion factor, first, the OD600 of a bacterial culture was measured. Second, a small volume (50-100 µL) of serial dilution of bacterial culture was spread plated on agar plates. After overnight culture, viable colonies on each agar plate were counted. Combined with plating volume and dilution factor, CFU/mL was calculated. Then OD600 measurement was correlated with CFU/mL measurement. The same correlation/conversion factor was used in all experiments.

**Preparation of CandyCollect devices for human subjects study**
For human subjects samples, CandyCollect devices were milled from 4 mm thick polystyrene sheets (Goodfellow, CAT# 725-602-13). Isomalt candy was applied to CandyCollect sticks in a kitchen following the hygiene guidance outlined in the Washington State Cottage Food Operations Law (RCW 69.22.040(2b-f(ii-iv)). Lab members who prepared CandyCollect were trained in food safety, had a Food Worker Card (WA State), and wore gloves and a mask during food preparation. The isomalt candy was prepared as described in our previous paper.[1,2,3] In brief, isomalt was gradually added to water, and food coloring was added with the last portion of isomalt. While cooling the isomalt candy, strawberry candy flavoring was quickly added to the mixture, and the isomalt was poured onto a marble slab to set. Then, the plasma treated CandyCollect polystyrene sticks were cleaned using hot water and dish soap. Desired portions of the isomalt candy were remelted and applied to the CandyCollect sticks using a silicone mold (Participants 1, 2 and 4, in Figure 5, used CandyCollects with 0.5 g candy and Participants 3, 5, and 6 used CandyCollects with 1.3 g candy). Then the CandyCollect devices were placed into polypropylene bags and heat sealed. Devices were stored in food preparation containers with a desiccant (Dry and Dry, Cat# B00DYKTS9C) until being sent to participants.

**Human subjects study details**

We conducted a clinical study at an ambulatory care clinic in Madison, Wisconsin, recruiting pediatric patients aged 5-17 years. The study was approved by the UW-Madison Health Sciences Institutional Review Board (#2021-1427). The details of recruitment eligibility, specimen collection, and laboratory analysis procedures were previously published.[3] In brief, upon testing positive for GAS by way of RADT performed on a pharyngeal swab as part of their clinical care, the assenting participant and their consenting caregiver were enrolled by the research nurse to complete the study after a clinical visit. A research nurse guided pediatric participants through collection of four oral specimen samples: two mouth swabs [COPAN ESwab™ (Becton, Dickinson and Company, Cat # R723482)] and two CandyCollect devices. When the participant completed sampling, the CandyCollect device was placed in a round-bottom, screwcap, Greiner Bio-One tube (Thermo Fisher Scientific Cat# 07-000-446), and the swabs in their respective tubes with ESwab™ buffer. Samples were stored in a -20 °C freezer for up to two weeks after which they were transferred on dry ice to a -80 °C freezer before being shipped to University of Washington for analysis.
